# Natural genetic diversity modulates structural stability changes caused by pathogenic variants in small proteins

**DOI:** 10.64898/2026.10.30.26366920

**Authors:** Tim Niemeijer, Morris A. Swertz, K. Joeri van der Velde

## Abstract

Clinical genomics relies heavily on linear reference genomes such as GRCh38, often ne-glecting natural sequence diversity that can modify variant expressivity and drive diagnostic health disparities. Meanwhile, variant interpretation is moving toward direct biophysical evidence of disrupted protein function, yet how natural genetic backgrounds modulate variant-induced protein stability changes remains uncharacterized. Here, we present a physics-based computational and data processing framework to evaluate how global sequence diversity alters the predicted thermodynamic impact of pathogenic mutations. Focusing on small human proteins (*≤* 100 residues), we reconstructed natural DNA haplotypes from 1000 Genomes Project data and introduced ClinVar pathogenic variants, yielding both natural and pathogenic protein haplotypes that were structurally modeled and energetically relaxed. Across 271 tested variants in 46 genes, structural stability changes (ΔΔ*G*) for 168 variants (62%) differed significantly depending on the underlying natural haplotype background (Kruskal-Wallis, Bonferroni-adjusted *p <* 0.01). These background-sensitive variants spanned 43 of the 46 genes (93%), suggesting that thermodynamic modulation by natural sequence context is a pervasive phenomenon across the human proteome. Accounting for haplotype context in protein stability profiling is therefore essential for equitable, ancestry-aware precision medicine.

## 1 Introduction

Modern precision medicine and clinical genomics rely on single linear reference genomes such as GRCh38 as the standard baseline for variant annotation and pathogenicity classification [1]. In reality, human population history has generated vast genetic diversity, with projects such as the 1000 Genomes Project (1kGP) showing that each individual carries a unique constellation of natural sequence variation [2]. Failing to account for this natural diversity in genetic context can lead to variant misclassification and exacerbate health disparities across diverse demographic groups [3, 4].

Furthermore, clinical variant classification is increasingly moving toward direct functional evidence rather than indirect associations of pathogenicity [5, 6, 7]. Central to this functional approach is the principle that a protein’s function depends on its thermodynamic stability, which must be maintained across diverse physiological environments [8, 9]. Recent methods have improved the prediction of protein stability, enabling large-scale assessments [10, 11].

Genetic background is known to modify the penetrance and phenotypic severity of disease-causing variation [12, 13], but how these modulating effects are shaped by populations haplo-types, particularly on a protein stability level, remains poorly understood [14, 15].

Here, we present a systematic framework to evaluate how natural haplotype backgrounds modulate structural stability changes (ΔΔ*G*) caused by pathogenic variants. To achieve high-resolution thermodynamic sampling, we focus on small proteins (*≤* 100 amino acids) and map disease-associated variants directly onto natural haplotypes derived from the 1kGP dataset. Using physics-based energy simulations, we demonstrate that natural sequence contexts systematically shift the calculated thermodynamic impact of pathogenic mutations, underscoring the importance of haplotype-aware stability prediction in clinical variant interpretation and precision medicine.

## 2 Results

We established a framework to reconstruct natural protein haplotypes from 1kGP data, map pathogenic ClinVar variants, model their 3D structures, and quantify thermodynamic stability via Rosetta free-energy scoring (Figure 1).

**Figure 1:**
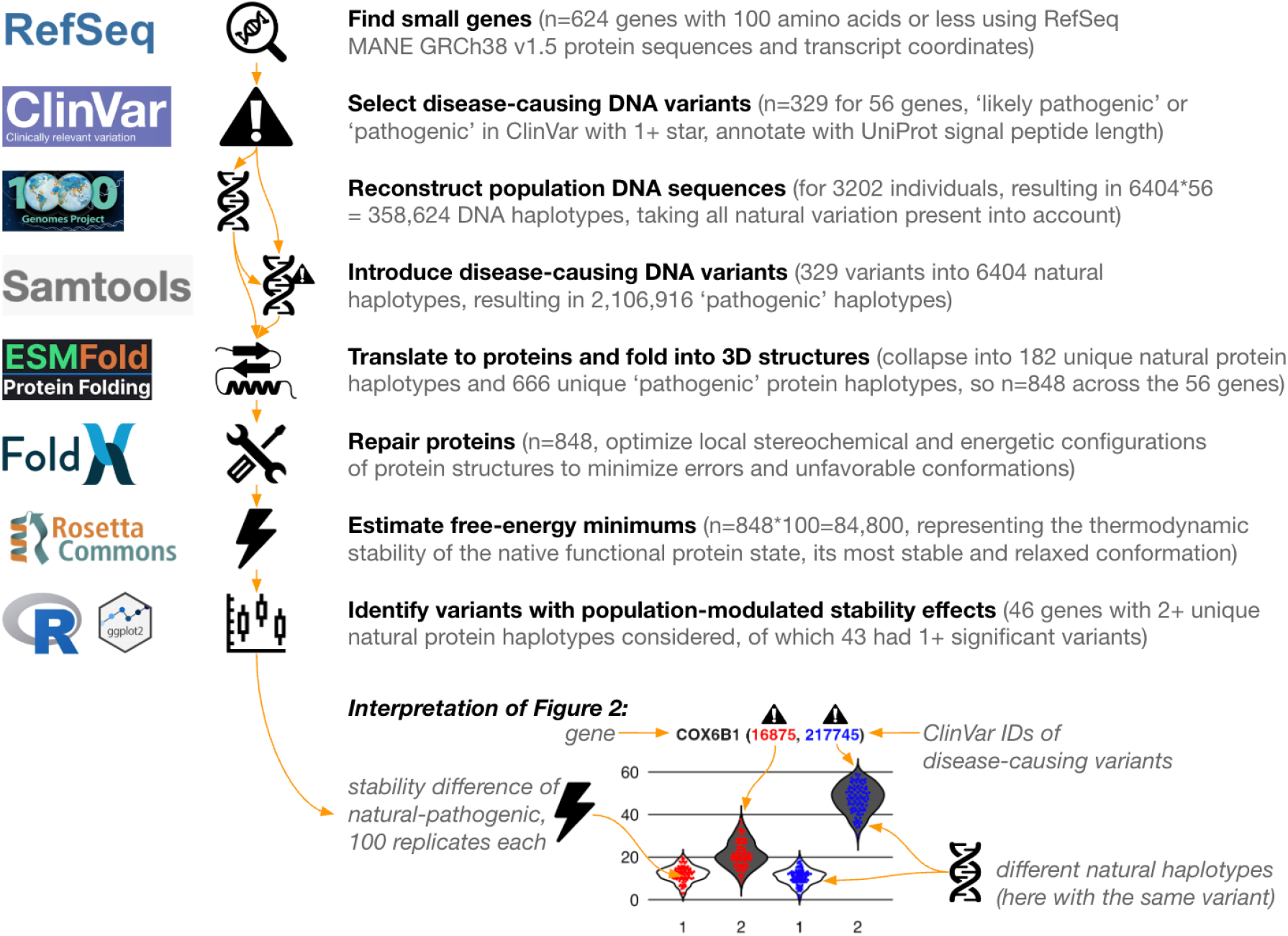
Overview of the framework. Integrated computational and data processing framework for reconstructing natural population haplotypes, modeling pathogenic variants, and executing physics-based energy calculations. The lower part is an interpretation guide for down-stream stability distributions in Figure 2. Icons from Noun Project, created by: Dawid Zyla, Ayub Irawan, Adrien Coquet, Kris.27, kusuma potter, UNKNOWN, and Vectors Point.

In total, we evaluated 271 pathogenic variants across 46 small proteins. Structural stability changes (ΔΔ*G*) induced by 168 variants (62%) differed significantly depending on the underlying natural haplotype background (Kruskal-Wallis test, Bonferroni adjusted *p <* 0.01, Figure 2). These background-sensitive variants were broadly distributed, spanning 43 of the 46 evaluated genes (93%). Full test results are shown in Table 1.

**Table 1:** Significance test results for haplotype-dependent thermodynamic stability. Each row displays variant-level Kruskal-Wallis test results evaluating the genetic background effect on stability changes caused by pathogenic variants in small proteins. Columns indicate gene, ClinVar variant ID, Bonferroni-adjusted *p*-value, and epsilon-squared effect size (*ϵ*^2^).

| Gene | ClinVar ID | Adj.*p*-val | Eff | Gene | ClinVar ID | Adj.*p*-val | Eff | Gene | ClinVar ID | Adj.*p*-val | Eff | Gene | ClinVar ID | Adj.*p*-val | Eff |
|---|---|---|---|---|---|---|---|---|---|---|---|---|---|---|---|
| MYMX | 1693533 | 5.54e-152 | 0.68 | HAMP | 4075845 | 2.95e-47 | 0.75 | DPM2 | 2745894 | 1.76e-19 | 0.32 | HAMP | 1350113 | 1.00e+00 | 0.01 |
| MYMX | 3747851 | 2.14e-151 | 0.68 | LOC128092249 | 2108844 | 8.12e-47 | 0.75 | DPM2 | 39446 | 2.27e-19 | 0.32 | HAMP | 2576337 | 1.00e+00 | 0.00 |
| PET100 | 2740392 | 5.24e-129 | 0.89 | CSTB | 3376548 | 1.36e-46 | 0.57 | MOCS2 | 2441783 | 7.70e-19 | 0.20 | HAMP | 4287 | 1.00e+00 | 0.00 |
| PET100 | 2848577 | 4.38e-121 | 0.84 | SMPX | 29948 | 3.78e-46 | 0.46 | GTF2H5 | 975159 | 9.74e-19 | 0.31 | LOC128092249 | 3702047 | 1.00e+00 | 0.00 |
| PDE6G | 1465534 | 4.62e-120 | 0.83 | SMPX | 3724097 | 5.86e-46 | 0.73 | DPM2 | 632533 | 2.98e-18 | 0.30 | LOC128092249 | 4845932 | 1.00e+00 | 0.00 |
| MYMX | 4845832 | 1.21e-119 | 0.54 | MOCS2 | 1236176 | 6.50e-46 | 0.45 | MOCS2 | 6111 | 4.15e-18 | 0.19 | MOCS2 | 1211302 | 1.00e+00 | 0.00 |
| SPINK1 | 1776795 | 7.01e-117 | 0.71 | NDUFB3 | 809137 | 7.04e-46 | 0.45 | PLN | 1709041 | 1.45e-17 | 0.42 | MOCS2 | 1406875 | 1.00e+00 | 0.00 |
| PDE6G | 1943178 | 4.62e-103 | 0.83 | SMPX | 3024082 | 1.05e-45 | 0.73 | SMPX | 1676666 | 1.64e-17 | 0.29 | MOCS2 | 1424517 | 1.00e+00 | 0.00 |
| PET100 | 1686017 | 3.67e-99 | 0.69 | PLN | 1072579 | 2.89e-45 | 0.72 | ATP5F1E | 30551 | 4.00e-17 | 0.22 | MOCS2 | 1458938 | 1.00e+00 | 0.00 |
| CSTA | 450304 | 3.09e-97 | 0.78 | PIGY | 222024 | 1.93e-44 | 0.71 | LYRM4 | 102450 | 6.56e-17 | 0.18 | MOCS2 | 1942085 | 1.00e+00 | 0.00 |
| PET100 | 128250 | 1.60e-88 | 0.62 | PRCD | 1929516 | 2.74e-44 | 0.71 | SMPX | 3601795 | 4.93e-14 | 0.24 | MOCS2 | 2071662 | 1.00e+00 | 0.00 |
| MOCS2 | 930940 | 2.05e-88 | 0.85 | PRCD | 987020 | 3.12e-44 | 0.71 | NDUFAF8 | 691641 | 6.69e-14 | 0.23 | MOCS2 | 2101843 | 1.00e+00 | 0.00 |
| MOCS2 | 2833050 | 5.20e-88 | 0.85 | PLN | 940239 | 6.55e-43 | 0.69 | PLN | 3706363 | 1.28e-13 | 0.33 | MOCS2 | 2108181 | 1.00e+00 | 0.00 |
| MYMX | 3747852 | 5.79e-88 | 0.40 | SMPX | 3347097 | 2.54e-42 | 0.52 | PLN | 3655536 | 1.75e-13 | 0.32 | MOCS2 | 2177297 | 1.00e+00 | 0.00 |
| PET100 | 125441 | 1.36e-87 | 0.61 | NDUFB3 | 1698952 | 3.09e-42 | 0.42 | SMPX | 1451817 | 2.02e-13 | 0.23 | MOCS2 | 2582660 | 1.00e+00 | 0.00 |
| PET100 | 3584209 | 1.36e-87 | 0.61 | SMPX | 29946 | 3.17e-42 | 0.67 | GTF2H5 | 2104 | 1.62e-12 | 0.21 | MOCS2 | 2696333 | 1.00e+00 | 0.00 |
| SPINK1 | 36780 | 9.06e-87 | 0.70 | DPM2 | 3707799 | 3.79e-42 | 0.67 | IER3IP1 | 30786 | 4.81e-12 | 0.13 | MOCS2 | 2727260 | 1.00e+00 | 0.00 |
| MOCS2 | 2153568 | 7.63e-83 | 0.80 | PRCD | 813201 | 5.32e-42 | 0.67 | PLN | 1977709 | 5.97e-12 | 0.29 | MOCS2 | 2745092 | 1.00e+00 | 0.00 |
| MOCS2 | 6113 | 4.31e-82 | 0.79 | PRCD | 37041 | 2.77e-41 | 0.66 | GTF2H5 | 2103 | 5.55e-11 | 0.19 | MOCS2 | 2747134 | 1.00e+00 | 0.00 |
| MOCS2 | 4711719 | 6.46e-82 | 0.79 | PRCD | 1379691 | 3.16e-41 | 0.66 | COXFA4 | 1176532 | 1.30e-10 | 0.26 | MOCS2 | 2750933 | 1.00e+00 | 0.00 |
| PET100 | 1458646 | 8.08e-78 | 0.55 | APOC3 | 17903 | 3.31e-41 | 0.66 | NDUFAF8 | 691643 | 6.45e-10 | 0.17 | MOCS2 | 2805758 | 1.00e+00 | 0.00 |
| SPINK1 | 1779356 | 1.06e-77 | 0.75 | COA5 | 31087 | 1.11e-40 | 0.65 | DPM3 | 585021 | 6.31e-09 | 0.22 | MOCS2 | 2826287 | 1.00e+00 | 0.00 |
| MOCS2 | 1454403 | 5.17e-77 | 0.74 | BBIP1 | 626905 | 6.54e-40 | 0.34 | GTF2H5 | 975158 | 6.59e-09 | 0.16 | MOCS2 | 2829748 | 1.00e+00 | 0.00 |
| SMPX | 1065047 | 3.01e-74 | 0.72 | HAMP | 4284 | 1.74e-39 | 0.63 | GTF2H5 | 4747548 | 2.13e-08 | 0.15 | MOCS2 | 2858763 | 1.00e+00 | 0.00 |
| NDUFAF8 | 691644 | 4.47e-74 | 0.89 | SMPX | 444056 | 1.01e-38 | 0.48 | DPM3 | 4702 | 2.28e-08 | 0.21 | MOCS2 | 2863141 | 1.00e+00 | 0.00 |
| LOC128092249 | 3346949 | 6.91e-73 | 0.87 | HAMP | 917401 | 3.12e-38 | 0.61 | PLN | 44580 | 4.76e-07 | 0.18 | MOCS2 | 2875294 | 1.00e+00 | 0.00 |
| MOCS2 | 452473 | 1.73e-71 | 0.69 | SMPX | 1676665 | 1.11e-37 | 0.60 | DPM3 | 694278 | 9.10e-07 | 0.17 | MOCS2 | 2902469 | 1.00e+00 | 0.00 |
| CSTB | 1455136 | 2.83e-70 | 0.68 | APOC3 | 17902 | 2.04e-35 | 0.57 | PLN | 1453340 | 1.32e-04 | 0.12 | MOCS2 | 2998110 | 1.00e+00 | 0.00 |
| MOCS2 | 1456543 | 2.99e-69 | 0.67 | PRCD | 143095 | 1.27e-33 | 0.54 | SPTSSA | 2505470 | 2.07e-04 | 0.12 | MOCS2 | 3592706 | 1.00e+00 | 0.00 |
| CSTB | 493324 | 9.80e-69 | 0.56 | BBIP1 | 126379 | 1.45e-33 | 0.29 | DPM3 | 1060967 | 4.94e-04 | 0.11 | MOCS2 | 3645068 | 1.00e+00 | 0.00 |
| SMPX | 975949 | 1.97e-68 | 0.56 | SPINK1 | 4688635 | 2.61e-32 | 0.33 | NDUFAF8 | 1690644 | 7.86e-04 | 0.08 | MOCS2 | 3774925 | 1.00e+00 | 0.00 |
| MOCS2 | 1236175 | 1.92e-66 | 0.65 | HRURF | 7342 | 5.15e-32 | 0.75 | APOA2 | 4541826 | 8.49e-03 | 0.08 | MOCS2 | 6108 | 1.00e+00 | 0.00 |
| SPINK1 | 1773780 | 7.51e-66 | 0.64 | SMPX | 445653 | 6.37e-32 | 0.40 | APOA2 | 2690543 | 2.61e-02 | 0.07 | MOCS2 | 6109 | 1.00e+00 | 0.00 |
| GON7 | 1321193 | 1.03e-64 | 0.63 | HRURF | 2001115 | 6.75e-32 | 0.75 | DPM3 | 658481 | 2.86e-02 | 0.07 | MOCS2 | 6110 | 1.00e+00 | 0.00 |
| UQCC3 | 161120 | 1.08e-64 | 0.46 | COX6B1 | 217745 | 6.84e-32 | 0.75 | DPM3 | 1989778 | 2.93e-02 | 0.07 | MOCS2 | 6115 | 1.00e+00 | 0.00 |
| CSTB | 8396 | 2.56e-64 | 0.77 | CKS1B | 1706624 | 6.84e-32 | 0.75 | DPM3 | 4736163 | 3.05e-02 | 0.07 | NDUFA2 | 7515 | 1.00e+00 | 0.00 |
| HAMP | 4285 | 8.36e-64 | 0.77 | COX14 | 214250 | 6.84e-32 | 0.75 | DPM3 | 2043355 | 3.11e-02 | 0.07 | NDUFAF8 | 691642 | 1.00e+00 | 0.00 |
| CSTB | 431700 | 1.55e-63 | 0.76 | HRURF | 2696995 | 8.37e-32 | 0.75 | COX14 | 31196 | 4.55e-02 | 0.07 | NDUFC1 | 1300558 | 1.00e+00 | 0.01 |
| GON7 | 1321194 | 3.29e-63 | 0.62 | CKS1B | 1706625 | 1.23e-30 | 0.72 | DPM3 | 694292 | 9.68e-02 | 0.06 | NDUFC1 | 2020770 | 1.00e+00 | 0.01 |
| NDUFB3 | 520909 | 3.36e-63 | 0.62 | COX7B | 39767 | 1.42e-30 | 0.72 | DPM3 | 374109 | 1.00e-01 | 0.06 | NDUFC1 | 2577991 | 1.00e+00 | 0.01 |
| MOCS2 | 2834251 | 4.01e-62 | 0.61 | HRURF | 830233 | 1.68e-30 | 0.72 | GTF2H5 | 975160 | 1.23e-01 | 0.05 | PDE6G | 13102 | 1.00e+00 | 0.00 |
| MOCS2 | 813890 | 4.01e-62 | 0.61 | COX7B | 39769 | 5.47e-30 | 0.70 | APOA2 | 17935 | 1.00e+00 | 0.00 | PDE6G | 3250311 | 1.00e+00 | 0.00 |
| GNRH1 | 156555 | 1.57e-61 | 0.74 | HRURF | 7343 | 5.82e-30 | 0.70 | CCL5 | 12740 | 1.00e+00 | 0.00 | PET100 | 2698099 | 1.00e+00 | 0.00 |
| HAMP | 4283 | 2.60e-61 | 0.74 | DPM2 | 2442789 | 3.48e-29 | 0.47 | COX7B | 39768 | 1.00e+00 | 0.00 | PET100 | 2788135 | 1.00e+00 | 0.00 |
| MOCS2 | 6116 | 9.76e-61 | 0.59 | HRURF | 1335706 | 9.17e-29 | 0.68 | COX7B | 449642 | 1.00e+00 | 0.00 | PET100 | 2889283 | 1.00e+00 | 0.00 |
| GNRH1 | 1388091 | 1.02e-60 | 0.73 | SMPX | 29945 | 1.73e-27 | 0.45 | COX8A | 222973 | 1.00e+00 | 0.00 | PET100 | 2967109 | 1.00e+00 | 0.00 |
| GNRH1 | 14417 | 1.02e-60 | 0.73 | PLN | 3720713 | 2.70e-27 | 0.64 | COXFA4 | 2444066 | 1.00e+00 | 0.00 | PIGY | 993284 | 1.00e+00 | 0.00 |
| CSTB | 55957 | 2.53e-60 | 0.73 | DPM2 | 1065328 | 8.63e-27 | 0.44 | COXFA4 | 3255019 | 1.00e+00 | 0.00 | PLN | 13637 | 1.00e+00 | 0.02 |
| MOCS2 | 1451432 | 3.64e-60 | 0.59 | UFM1 | 559446 | 1.75e-26 | 0.62 | COXFA4 | 3898851 | 1.00e+00 | 0.00 | PRCD | 1184568 | 1.00e+00 | 0.00 |
| MOCS2 | 587508 | 1.30e-58 | 0.57 | UFM1 | 2428600 | 4.33e-26 | 0.62 | COXFA4 | 977513 | 1.00e+00 | 0.00 | PRCD | 1343514 | 1.00e+00 | 0.00 |
| NDUFA2 | 214711 | 4.83e-58 | 0.57 | NDUFA1 | 11649 | 5.69e-26 | 0.61 | CSTA | 208472 | 1.00e+00 | 0.00 | PRCD | 1427185 | 1.00e+00 | 0.00 |
| TMEM167A | 4848193 | 5.48e-56 | 0.89 | MOCS2 | 6112 | 8.14e-26 | 0.26 | CSTA | 208473 | 1.00e+00 | 0.00 | PRCD | 1976161 | 1.00e+00 | 0.00 |
| CSTB | 161418 | 1.14e-55 | 0.67 | HRURF | 7341 | 1.17e-25 | 0.60 | CSTA | 29892 | 1.00e+00 | 0.02 | PRCD | 3583072 | 1.00e+00 | 0.00 |
| NDUFB3 | 1678606 | 2.02e-53 | 0.52 | SMPX | 1676664 | 1.27e-25 | 0.42 | CSTA | 29893 | 1.00e+00 | 0.00 | PRCD | 3637386 | 1.00e+00 | 0.00 |
| NDUFB3 | 252575 | 5.10e-53 | 0.52 | DPM3 | 2762867 | 6.09e-25 | 0.41 | CSTA | 3357701 | 1.00e+00 | 0.00 | PRCD | 866216 | 1.00e+00 | 0.00 |
| SMPX | 40063 | 1.05e-52 | 0.44 | COX6A2 | 638271 | 6.50e-25 | 0.31 | CSTA | 3779552 | 1.00e+00 | 0.00 | SMPX | 1297079 | 1.00e+00 | 0.00 |
| CSTB | 659184 | 8.21e-52 | 0.63 | NDUFA2 | 978071 | 1.02e-24 | 0.26 | CSTB | 4725737 | 1.00e+00 | 0.00 | SMPX | 3601797 | 1.00e+00 | 0.00 |
| CSTB | 55960 | 1.50e-51 | 0.62 | SMPX | 29947 | 1.92e-24 | 0.40 | CSTB | 55958 | 1.00e+00 | 0.00 | SMPX | 3677196 | 1.00e+00 | 0.00 |
| CSTB | 1791620 | 2.48e-51 | 0.62 | HRURF | 7344 | 2.88e-24 | 0.57 | CSTB | 55959 | 1.00e+00 | 0.00 | SMPX | 417903 | 1.00e+00 | 0.00 |
| CSTB | 8397 | 6.95e-51 | 0.62 | SMPX | 1445549 | 7.79e-24 | 0.39 | CSTB | 55961 | 1.00e+00 | 0.00 | SPINK1 | 132142 | 1.00e+00 | 0.00 |
| MOCS2 | 6114 | 1.21e-50 | 0.50 | NDUFA1 | 11648 | 9.63e-24 | 0.56 | CSTB | 8395 | 1.00e+00 | 0.00 | SPINK1 | 13761 | 1.00e+00 | 0.00 |
| NOP10 | 2506513 | 2.23e-50 | 0.80 | PLN | 13636 | 1.95e-23 | 0.55 | DPM2 | 39447 | 1.00e+00 | 0.00 | SPINK1 | 13764 | 1.00e+00 | 0.00 |
| FXYD2 | 7684 | 5.39e-50 | 0.61 | COX6B1 | 16875 | 8.14e-22 | 0.52 | DPM3 | 1074098 | 1.00e+00 | 0.00 | SPINK1 | 1747977 | 1.00e+00 | 0.00 |
| CSTB | 1807163 | 1.86e-49 | 0.60 | HRURF | 3646769 | 8.55e-21 | 0.49 | DPM3 | 1446016 | 1.00e+00 | 0.02 | SPINK1 | 547787 | 1.00e+00 | 0.00 |
| SMPX | 987023 | 5.54e-49 | 0.78 | DPM2 | 2802571 | 1.63e-20 | 0.34 | GTF2H5 | 1382813 | 1.00e+00 | 0.00 | SPINK1 | 633006 | 1.00e+00 | 0.00 |
| CSTB | 575156 | 8.52e-48 | 0.58 | MOCS2 | 2021553 | 1.94e-20 | 0.21 | GTF2H5 | 3689291 | 1.00e+00 | 0.00 | SPTSSA | 2505468 | 1.00e+00 | 0.00 |
| CSTB | 8400 | 9.41e-48 | 0.58 | DPM2 | 1065329 | 2.86e-20 | 0.33 | GTF2H5 | 587425 | 1.00e+00 | 0.00 | UFM1 | 3358901 | 1.00e+00 | 0.00 |
| CSTB | 809294 | 1.53e-47 | 0.58 | PET117 | 981504 | 7.17e-20 | 0.21 | GTF2H5 | 2136487 | 1.00e+00 | 0.01 |  |  |  |  |

**Figure 2:**
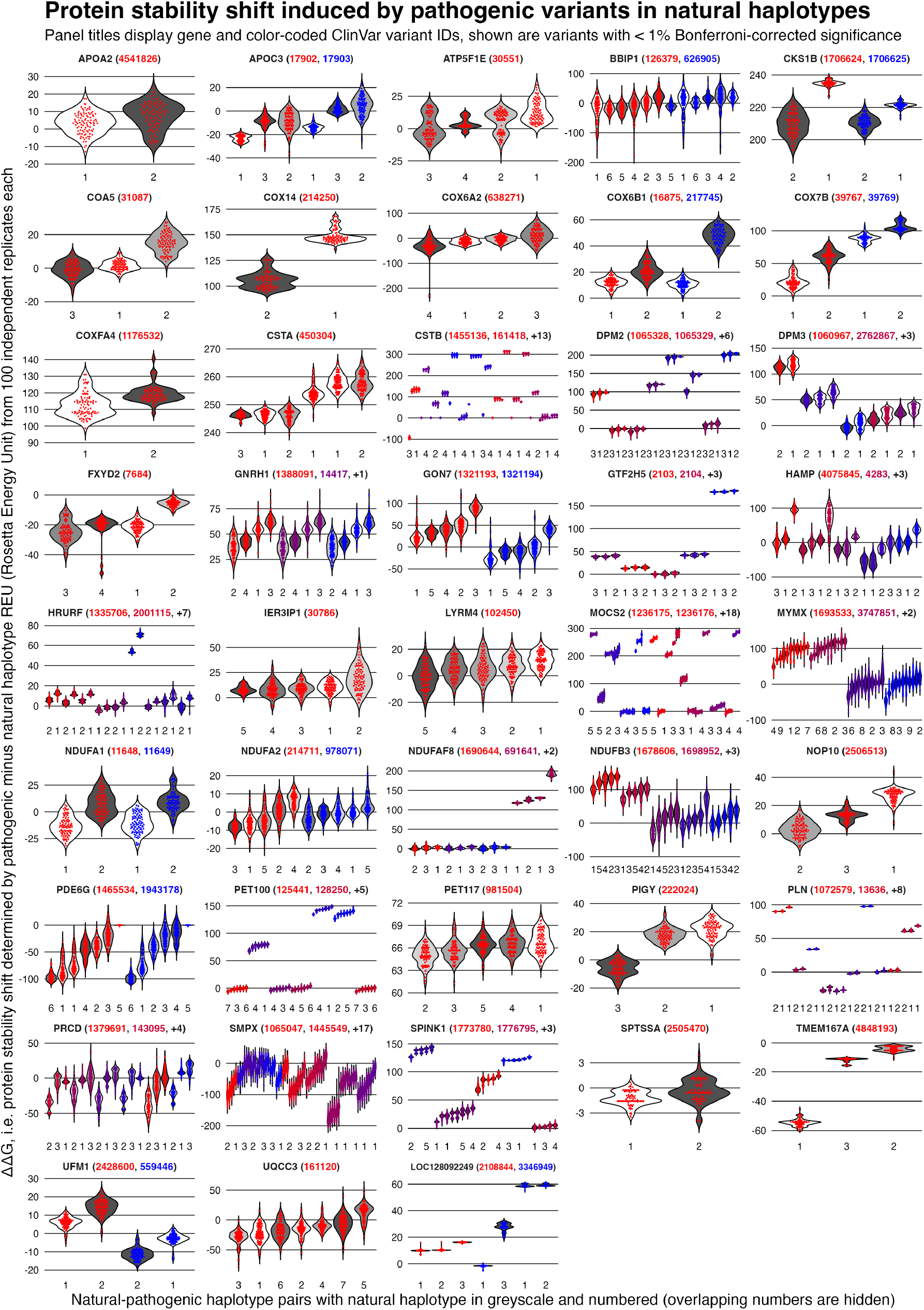
Modulation of pathogenic variant stability across natural protein haplotypes. Panel titles show gene names and ClinVar IDs of significant variants. Violins depict structural stability changes (ΔΔ*G*) across 100 replicates per natural-pathogenic haplotype pair. Natural background haplotypes are shown in greyscale and numbered along the x-axis

To contextualize the demographic origin of the background-dependent stability shifts, we stratified the analysed natural protein haplotypes across global superpopulations (Figure 3). While many protein haplotypes are widely shared across diverse human populations, others appear to be ancestry-specific, though sample counts vary greatly, where major haplotypes may represent thousands of individuals alongside rare alternative haplotypes.

**Figure 3:**
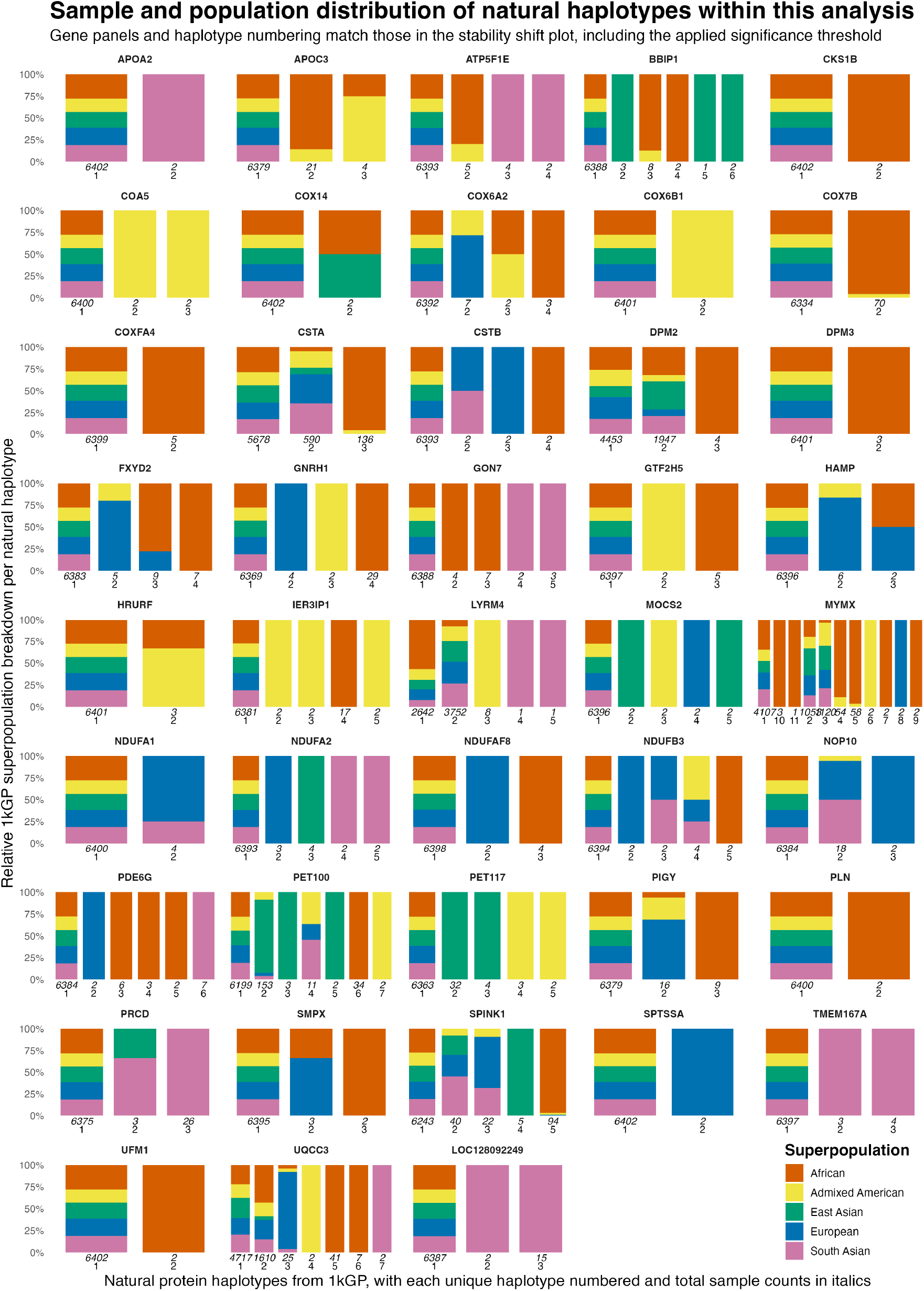
Population distribution of natural protein haplotypes. Each panel represents a gene, with natural protein haplotypes numbered and their total sample counts shown in italics. The y-axis depicts the relative proportion of each haplotype across 1000 Genomes Project superpopulations. Genes and haplotypes correspond directly to Figure 2.

## 3 Conclusion

Our findings demonstrate that naturally occurring protein sequence diversity can substantially influence the predicted structural energy changes associated with pathogenic variants. Across 271 variants in 46 human proteins, we identified significant haplotype-dependent differences for 168 variants, highlighting the importance of considering natural sequence backgrounds when assessing the potential structural consequences of pathogenic variation.

## 4 Discussion

By bridging population and clinical genetics with high-resolution biophysical modeling, our findings show that variant pathogenicity cannot be decoupled from genetic background. Accounting for population-specific haplotype contexts in functional protein analyses are expected to refine our understanding of variable expressivity and penetrance, offering a vital step forward for equitable, ancestry-aware precision medicine and accurate clinical variant annotation.

Nevertheless, key limitations must be acknowledged. First, our analysis highlights the challenge of uneven representation of natural haplotypes across the sampled population. Our observed natural haplotype frequencies exhibit a skewed distribution, with sample counts spanning from single digits up to thousands. This uneven representation may limit the characterisation of rare haplotypes and the generalisability of our findings across populations. Consequently, advancing haplotype-aware precision medicine will require sequencing efforts that prioritise global population diversity and the inclusion of under-represented genetic backgrounds, rather than focusing solely on increasing overall sample size.

Furthermore, although restricting our initial scope to small single-domain proteins enabled extensive energy landscape sampling, extending this framework to larger and more complex proteins will require substantial methodological and computational advances to account for long-range allosteric effects, conformational dynamics, ligand binding, protein-protein interactions, and the energetics of quaternary structure interfaces. Incorporating disease-relevant and tissue-specific isoforms beyond canonical MANE Select transcripts would further improve biological relevance[16]. Yet, even at this heightened level of biological representation, measurable molecular vulnerabilities may not be direct determinants of disease expressivity, as downstream phenotypes remain ultimately governed by cellular homeostatic networks, compensatory pathways, and environmental factors. Given that individuals typically carry two haplotypes for each gene, combinations of protein variants across interacting partners create an increasingly complex interaction landscape. This underscores the need for personalised proteogenomics to understand how individual genetic variation shapes cellular function and may ultimately contribute to differences in clinical outcomes.

Finally, numerous studies have documented associations between genetic background and variable expressivity across the significant genes identified in our analysis. Notable examples include (i) the broad spectrum of SPINK1 disease penetrance, which reflects the interplay between causal variants and genetic background [17], (ii) ancestry-modulated plasma triglyceride reductions observed in heterozygous APOC3 loss-of-function carriers [18], and (iii) population-specific CCL5 variations influencing HIV-1 susceptibility and tuberculosis risk [19, 20, 21]. Future research might investigate the biophysical mechanisms underlying these background-dependent effects and evaluate their functional impact in vivo, though moving beyond these initial boundaries will require long-read, phased, and integrative VCF datasets to resolve complex cis-regulatory haplotype structures, scaling our framework to complex protein dynamics, and incorporating broader global genomic diversity.

## 5 Methods

The computational and data processing framework developed in this study consists of data selection, haplotype reconstruction, structural modeling, and thermodynamic estimation as outlined in Figure 1. The subsections of here correspond directly to each step of the outlined framework, detailing the precise parameters, tools, and mathematical criteria applied. The concrete implementation can be found in the “small-gene-bulk-analysis folder” of the source code, consisting of various scripts and orchestrators: 0-all-small-genes.sh, 1-extract-small-genevariants-from-clinvar.sh, 2-signal-peptide-lengths.sh, 3-reconstruct-1kg-patho-haplotypes.sh, 4-fold.sh, 5-repair.sh, 6-relax.sh, 7-lowest.sh, and 8-analysis.R. All source code and key results are available at https://github.com/molgenis/MORPHA and permanently archived at https://zenodo.org/records/23727210.

### 5.1 Find small genes

To construct the dataset of small proteins, reference annotation and sequence data were retrieved from the Matched Annotation from NCBI and EBI (MANE) project release 1.5 for GRCh38. Specifically, human reference protein sequences (MANE.GRCh38.v1.5.refseq_protein.faa.gz) and the corresponding metadata summary table (MANE.GRCh38.v1.5.summary.txt.gz) were acquired from the NCBI FTP repository.

Amino acid sequence lengths for all proteins were parsed directly from the FASTA records. The proteins were cross-referenced with the MANE summary metadata and filtered for transcripts encoding products with an amino acid length of 100 residues or fewer. This step resulted in a list of 624 genes and includes gene symbols, RefSeq protein identifiers (NP_), RefSeq transcript identifiers (NM_), and sequence lengths.

### 5.2 Select disease-causing DNA variants

To capture high-confidence pathogenic variation for the 624 small proteins, variant records were queried from ClinVar [22] (version 20260503) using bcftools [23]. Strict filtering criteria were applied to isolate clinically relevant alterations. Clinical significance (CLNSIG) was re-stricted to variants explicitly annotated as Pathogenic, Likely_pathogenic, or Pathogenic/Likely_pathogenic. Review status (CLNREVSTAT) excluded entries lacking review assertions. Variants of extreme impact such as whole exon and gene deletions were excluded by restricting both reference and alternate allele lengths to 50 base pairs or fewer. Genomic coordinates for each target transcript were mapped using MANE RefSeq GFF annotation (MANE.GRCh38.v1.5.refseq_genomic.gff), with automatic harmonization of chromosome names between ClinVar and GFF sources. For each gene, absolute hg38 genomic coordinates were converted to transcript-relative positions (pos=hg38pos-start+1). The resulting filtered variants were compiled into gene-specific, compressed, and indexed VCF files (variants.vcf.gz) via bgzip and tabix. This resulted in 329 pathogenic variants for 56 genes.

To obtain signal peptide annotations for these 56 genes, automated queries were executed against the UniProt [24] Knowledgebase (UniProtKB) REST API at https://rest.uniprot.org. For each gene, the RefSeq protein identifier was mapped against reviewed human entries (organism_id:9606, reviewed:true) using explicit cross-reference parameters (database:RefSeq). Full-entry JSON records were programmatically retrieved for each protein accession. Feature annotation arrays were parsed using jq to extract structural topology metrics, specifically identifying regions annotated with the “Signal” feature type. Coordinate boundaries for predicted signal peptides were captured to compute the total span (signal length = signal end − signal start + 1). Processed records, including tracking states for successful mappings and edge-case exceptions were compiled into a structured tracking table. Signal peptides were present in APOA2, APOC3, GNRH1, CCL5, SPINK1, HAMP and TMEM167A. For LOC128092249 there was no UniProt match.

### 5.3 Reconstruct population DNA sequences

The shell script genename2uniqproteinhaplo.sh constructs individual-specific protein haplotypes from 1kGP data. The GRCh38 reference genome, MANE GFF annotations, and 1kGP phased VCF files were automatically downloaded for the target genomic region. An preprocessing function was implemented to parse and expand complex SVs denoted by symbolic alleles (deletions, duplications, and insertions such as Alu, SVA, and LINE1s) into the DNA sequence representations. VCF files were expanded and normalized accordingly by bcftools to fit all variant notations to their matching positions. For every individual and haplotype (h1/h2) across the 1kGP cohort, local consensus haplotype sequences were generated using bcftools consensus mapped against the gene region. Custom chain files and the helper script project_gff_by_chain_preserve_features.py performed coordinate liftover of gene GFF annotations onto each sample’s individual coordinate space. CDS sequences were then translated to proteins using gffread [25]. Reconstructed protein sequences were truncated at premature stop codons (*). To model the impact of mutations in a mature protein context, precursor signal peptide sequences were removed based on their UniProt signal peptide annotations. In an edge case of a stop codon residing inside a signal peptide, what was left of the signal peptide was retained to keep a consistent n=6404 in the overall analysis. Identical sequences were clustered into unique natural protein haplotypes with their corresponding sample and haplotype identifiers (e.g. HG01488.h2, HG01704.h1, etc).

### 5.4 Introduce disease-causing DNA variants

The shell script genename2pathohaplo.sh systematically introduced pathogenic variation onto individual 1kGP haplotypes. It validated input variant identifiers in the VCF for uniqueness and checked that CDS records in the gene-specific MANE GFF contain only the supplied protein ID and no other. For each variant and individual sample haplotype, chain files were localized, and variant coordinates were lifted over to sample-specific space using CrossMap vcf[26]. Variant redundancy was handled in case the allele already exists in the haplotype, otherwise it applies valid mutations via bcftools consensus to generate mutant FASTA sequences. GFF annotations were lifted over to the mutant coordinate space, followed by gffread translation into proteins. Processed mutant sequences were truncated at stop-gains, trimmed for signal peptides, and aggregated into unique pathogenic protein haplotype files.

It should be noted that identical natural backgrounds can diverge into multiple pathogenic protein haplotypes, whereas distinct natural backgrounds can converge into the same pathogenic haplotype. For instance, a frameshift variant in CSTA (ClinVar ID 450304) yields three distinct pathogenic protein haplotypes from a single natural protein haplotype due to DNA-level sequence variation. Conversely, introducing this variant across three distinct natural backgrounds yields an identical pathogenic protein haplotype because the resulting truncation at residue 42 removes downstream natural variation near the C-terminus (positions 91 and 96 of 98).

### 5.5 Translate to proteins and fold into 3D structures

To determine the three-dimensional structures of all curated natural and pathogenic protein variants, an automated structural prediction wrapper script (genename2foldedhaplo.sh) was used. The script systematically verified the presence of both natural and pathogenic variant haplotype FASTA files across all target genes, stopping with an error log if any required sequence files were absent. For validated genes, FASTA records were parsed using structured text-processing workflows (awk) to extract unique variant identifiers and remove formatting line breaks. Structural models were generated by submitting individual amino acid sequences via HTTP POST requests to a local structure prediction server endpoint running ESMFold [27], source code available at https://github.com/joerivandervelde/esmfold-server. Resulting atomic coordinates were stored as standard PDB files.

### 5.6 Repair proteins

Header identifiers were parsed from both natural and pathogenic FASTA files to establish a definitive inventory of expected protein structure PDB files which was cross-referenced against the physical files. Protein structures were subsequently subjected to structural optimization using the RepairPDB command of FoldX 5.1 [28]. RepairPDB identifies and resolves bad atomic contacts, steric overlaps, and unfavorable side-chain torsion angles by evaluating local rotamer combinations against the FoldX empirical force field. This initial cleanup removes severe van der Waals clashes and optimizes local hydrogen-bonding networks without altering the global tertiary fold. By mitigating localized energy spikes, FoldX provides a well-behaved starting structure, preventing the subsequent Rosetta’s FastRelax protocol from getting trapped in high-energy local minima or distorting the backbone during its iterative repacking and gradient-based backbone/side-chain relaxation sweeps.

### 5.7 Estimate free-energy minimums

To compute robust Gibbs free energy change (Δ*G*) for all natural and pathogenic protein haplotypes, high-throughput structural relaxation was performed using the FastRelax protocol in the Rosetta software suite (version 3.15) [29, 30, 31] with the ref2015 scoring function, out-putting results in Rosetta Energy Units (REU). While REUs are dimensionless and not directly calibrated to physical units like kcal/mol, Rosettas score functions are parameterized against experimental protein stabilities such that lower REU values correlate with more favorable folding thermodynamics. Furthermore, because FoldX uses empirical energy functions optimized for relative single-point mutations (ΔΔ*G*) via error cancellation rather than absolute free-energy estimation (Δ*G*), Rosetta FastRelax was selected to independently evaluate absolute conformational energies across diverse protein backbones.

To thoroughly sample the local conformational landscape, FastRelax was executed with nstruct=100 independent trajectories per protein structure. Generating 100 independent replicates allows the stochastic Monte Carlo minimization and side-chain repacking sweeps to escape shallow local traps, yielding an ensemble of diverse, low-energy structural conformations. Individual intermediate trajectory PDB and score files were automatically pruned, retaining only the total Δ*G* estimate from each trajectory for downstream analysis assembled into a single commadelimited matrix per haplotype. As part of quality-control it was noticed that the FoldX-repaired FXYD2 protein variant 1’ structure caused unphysical energy spikes during Rosetta evaluation, leading us to fall back on the original ESMFold-predicted (unrepaired) structure.

### 5.8 Identify variants with population-modulated stability effects

Statistical data analysis was done in R version 4.6.0. The R script 8-analysis.R analysed the impact of pathogenic genetic variants on protein stability across naturally occurring protein haplotypes. It combined variant data, protein sequences, and Rosetta energy minimisation results for a set of target genes, linking pathogenic haplotypes to their corresponding natural haplotypes through their introduced pathogenic variation in individuals from the 1000 Genomes Project (1kGP). A total of 10 genes without any natural protein haplotype variation within the 1kGP cohort were dropped (TIMM8A, GNG3, RPS21, RPS27, RPS29, SNRPE, IQSEC2, TOMM7, ATP5MK and SVBP), leaving 46 genes and 271 pathogenic variants for further analysis. For each pathogenic variant it calculated the change in predicted protein stability changes (ΔΔ*G*) by comparing the energy of the pathogenic haplotype with that of its natural counterpart across 100 replicates. Kruskal-Wallis tests with Bonferroni-adjusted *p <* 0.01 were then used to identify variants whose stability effects differed significantly between pairs of natural haplotype backgrounds and derived pathogenic haplotypes. Summary statistics show the proportion of significant variants and the percentage of genes containing at least one significant pathogenic variant. Data visualization was performed with the ggplot2 package, a multi-panel violin plot showing stability shifts for significant variants and a proportional stacked bar chart illustrating the population distribution of the corresponding natural haplotypes across the five 1kGP superpopulations.

## Declarations

### Ethics approval and consent to participate

Not applicable.

### Consent for publication

Not applicable.

### Availability of data and materials

All used data is publicly available. 1kGP vcf and tbi files were downloaded from https://ftp.1000genomes.ebi.ac.uk/vol1/ftp/data_collections/1000G_2504_high_coverage/working/20220422_3202_phased_SNV_INDEL_SV, MANE RefSeq gff, faa and summary from https://ftp.ncbi.nlm.nih.gov/refseq/MANE/MANE_human/release_1.5/, ClinVar vcf and tbi at https://ftp.ncbi.nlm.nih.gov/pub/clinvar/vcf_GRCh38/archive_2.0/2026/clinvar_20260503.vcf.gz, GRCh38 fa and fa.fai from https://ftp.1000genomes.ebi.ac.uk/vol1/ftp/technical/reference/GRCh38_reference_genome/. All generated data and results of this project are publicly available at https://github.com/molgenis/MORPHA and https://zenodo.org/records/23727210.

### Availability of source code

All code is available as free and open source software under the GNU Lesser General Public License (LGPL) v3.0 at https://github.com/molgenis/MORPHA.

## Data Availability

All generated data and results of this project are publicly available at https://github.com/molgenis/MORPHA and https://zenodo.org/records/23727210.

https://zenodo.org/records/23727210

## Author contributions

T.N., K.J.V. conceived the project. K.J.V. performed the data processing. T.N. and K.J.V. wrote the manuscript with critical input and revisions from M.A.S. All authors have reviewed and approved the manuscript.

## Acknowledgements

We thank the 1000 Genomes Project for providing open access to global genomic variation data. Special thanks are owed to the study participants from diverse populations around the world whose voluntary participation laid the foundation for this work. We would like to thank our colleagues at Genomics Coordination Center for engineering support and access to the UMCG high performance compute cluster.

## References

[1] Mireia Costa, Alberto García S, Ana León, and Oscar Pastor. The promises and pitfalls of automated variant interpretation: a comprehensive review. Briefings in Bioinformatics, 26(5), August 2025.

[2] Marta Byrska-Bishop, Uday S. Evani, Xuefang Zhao, et al. High-coverage whole-genome sequencing of the expanded 1000 genomes project cohort including 602 trios. Cell, 185(18):34263440.e19, 2022.

[3] Arjun K. Manrai, Birgit H. Funke, Heidi L. Rehm, Morten S. Olesen, Bradley A. Maron, Peter Szolovits, David M. Margulies, Joseph Loscalzo, and Isaac S. Kohane. Genetic misdiagnoses and the potential for health disparities. New England Journal of Medicine, 375(7):655665, August 2016.

[4] Rachel Hoffing, Daniel Zeiberg, Sarah L. Stenton, Matthew Mort, David N. Cooper, Matthew W. Hahn, Anne ODonnell-Luria, Lucas D. Ward, and Predrag Radivojac. Ancestry-specific performance of variant effect predictors in clinical variant classification. bioRxiv [Preprint], February 2026.

[5] Anupam Banerjee, Anthony T. Bogetti, and Ivet Bahar. Accurate identification and mechanistic evaluation of pathogenic missense variants with rhapsody-2. Proceedings of the National Academy of Sciences, 122(18), May 2025.

[6] Matteo Arnaudi, Mattia Utichi, Kristine Degn, et al. Mavisp: A modular structurebased framework for protein variant effects. Protein Science, 35(5), April 2026.

[7] Tim Niemeijer, René Mulder Helga Westers, Jan D. H. Jongbloed, Bart Charbon, Birgit Sikkema-Raddatz, Lennart F. Johansson, Mariëlle E. van Gijn, Cleo C. van Diemen, Dennis Hendriksen, Kristin M. Abbott, Willem T. K. Maassen, Morris A. Swertz, and K. Joeri van der Velde. Dave: how to use explainable ai to interpret missense variants for genome diagnostics based on functional protein modeling. F1000Research, 15:1345, August 2026.

[8] Nobuhiko Tokuriki and Dan S Tawfik. Stability effects of mutations and protein evolvability. Current Opinion in Structural Biology, 19(5):596604, October 2009.

[9] Tiziana Sanavia, Giovanni Birolo, Ludovica Montanucci, Paola Turina, Emidio Capriotti, and Piero Fariselli. Limitations and challenges in protein stability prediction upon genome variations: towards future applications in precision medicine. Computational and Structural Biotechnology Journal, 18:19681979, January 2020.

[10] Rebecca F. Alford, Andrew Leaver-Fay, Jeliazko R. Jeliazkov, Matthew J. OMeara, Frank P. DiMaio, Hahnbeom Park, Maxim V. Shapovalov, P. Douglas Renfrew, Vikram K. Mulligan, Kalli Kappel, Jason W. Labonte, Michael S. Pacella, Richard Bonneau, Philip Bradley, Roland L. Dunbrack, Rhiju Das, David Baker, Brian Kuhlman, Tanja Kortemme, and Jeffrey J. Gray. The rosetta all-atom energy function for macromolecular modeling and design. Journal of Chemical Theory and Computation, 13(6):30313048, May 2017.

[11] Valentina Sora, Adrian Otamendi Laspiur, Kristine Degn, Matteo Arnaudi, Mattia Utichi, Ludovica Beltrame, Dayana De Menezes, Matteo Orlandi, Ulrik Kristoffer Stoltze, Olga Rigina, Peter Wad Sackett, Karin Wadt, Kjeld Schmiegelow, Matteo Tiberti, and Elena Papaleo. Rosettaddgprediction for highthroughput mutational scans: From stability to binding. Protein Science, 32(1), December 2022.

[12] Tali Gidalevitz, Thomas Krupinski, Susana Garcia, and Richard I. Morimoto. Destabilizing protein polymorphisms in the genetic background direct phenotypic expression of mutant sod1 toxicity. PLoS Genetics, 5(3):e1000399, March 2009.

[13] Akl C. Fahed, Minxian Wang, Julian R. Homburger, Aniruddh P. Patel, Alexander G. Bick, Cynthia L. Neben, Carmen Lai, Deanna Brockman, Anthony Philippakis, Patrick T. Ellinor, Christopher A. Cassa, Matthew Lebo, Kenney Ng, Eric S. Lander, Alicia Y. Zhou, Sekar Kathiresan, and Amit V. Khera. Polygenic background modifies penetrance of monogenic variants for tier 1 genomic conditions. Nature Communications, 11(1), August 2020.

[14] Konrad J. Karczewski, Laurent C. Francioli, Grace Tiao, et al. The mutational constraint spectrum quantified from variation in 141,456 humans. Nature, 581(7809):434443, May 2020.

[15] Angela Wei, Richard Border, Boyang Fu, Sinéad Cullina, Nadav Brandes, Seon-Kyeong Jang, Sriram Sankararaman, Eimear E. Kenny, Miriam S. Udler, Vasilis Ntranos, Noah Zaitlen, and Valerie A. Arboleda. Investigating the sources of variable impact of pathogenic variants in monogenic metabolic conditions. Nature Communications, 16(1), 2025.

[16] Giovanna Weykopf, Mihaly Badonyi, Elias T. Friman, et al. Disease-associated genetic variants can cause missense effects in tissue-specific protein isoforms. Nature Communications, 17(1), 2026.

[17] Yuan-Chen Wang, Emmanuelle Masson, Qi-Wen Wang, Emmanuelle Génin, Gérald Le Gac, Yann Fichou, David N. Cooper, Zhuan Liao, Claude Férec, Wen-Bin Zou, and Jian-Min Chen. Spink1-related chronic pancreatitis: A model that encapsulates the spectrum of variant effects, genetic complexity, and classificatory challenges. The American Journal of Human Genetics, 112(9):20432066, 2025.

[18] The TG and HDL Working Group of the Exome Sequencing Project, National Heart, Lung, and Blood Institute. Loss-of-function mutations in apoc3, triglycerides, and coronary disease. New England Journal of Medicine, 371(1):2231, 2014.

[19] Qinghua Liao, Jianglong He, Xiaoquan Li, Jialin Tang, Tinli Jin, and Guoliang Hu. Association between chemotactic chemokine ligand 5-403g/a polymorphism and risk of human immunodeficiency virus-1 infection: a meta-analysis. OncoTargets and Therapy, page 727, April 2015.

[20] Marcos Jessé Abrahão Silva, Rebecca Lobato Marinho, Pabllo Antonny Silva dos Santos, Carolynne Silva dos Santos, Layana Rufino Ribeiro, Yan Corrêa Rodrigues, Karla Valéria Batista Lima, and Luana Nepomuceno Gondim Costa Lima. The association between ccl5/rantes snps and susceptibility to hiv-1 infection: A meta-analysis. Viruses, 15(9):1958, 2023.

[21] Yun-Feng Sheng and Qi Qi. Association of chemotactic chemokine ligand 5rs2107538polymorphism with tuberculosis susceptibility: A meta-analysis. Innate Immunity, 26(5):358363, December 2019.

[22] Melissa J. Landrum, Jennifer M. Lee, George R. Riley, Wonhee Jang, Wendy S. Rubinstein, Deanna M. Church, and Donna R. Maglott. Clinvar: public archive of relationships among sequence variation and human phenotype. Nucleic Acids Research, 42(D1):D980D985, November 2013.

[23] Petr Danecek, James K Bonfield, Jennifer Liddle, John Marshall, Valeriu Ohan, Martin O Pollard, Andrew Whitwham, Thomas Keane, Shane A McCarthy, Robert M Davies, and Heng Li. Twelve years of samtools and bcftools. GigaScience, 10(2), January 2021.

[24] Alex Bateman, Maria-Jesus Martin, Sandra Orchard, et al. Uniprot: the universal protein knowledgebase in 2023. Nucleic Acids Research, 51(D1):D523D531, November 2022.

[25] Geo Pertea and Mihaela Pertea. Gff utilities: Gffread and gffcompare. F1000Research, 9:304, 2020.

[26] Hao Zhao, Zhifu Sun, Jing Wang, Haojie Huang, Jean-Pierre Kocher, and Liguo Wang. Crossmap: a versatile tool for coordinate conversion between genome assemblies. Bioinformatics, 30(7):10061007, December 2013.

[27] Zeming Lin, Halil Akin, Roshan Rao, et al. Evolutionary-scale prediction of atomic-level protein structure with a language model. Science, 379(6637):11231130, March 2023.

[28] Javier Delgado, Raul Reche, Damiano Cianferoni, Gabriele Orlando, Rob van der Kant, Frederic Rousseau, Joost Schymkowitz, and Luis Serrano. Foldx force field revisited, an improved version. Bioinformatics, 41(2), February 2025.

[29] Michael D. Tyka, Daniel A. Keedy, Ingemar André, Frank DiMaio, Yifan Song, David C. Richardson, Jane S. Richardson, and David Baker. Alternate states of proteins revealed by detailed energy landscape mapping. Journal of Molecular Biology, 405(2):607618, January 2011.

[30] Firas Khatib, Seth Cooper, Michael D. Tyka, Kefan Xu, Ilya Makedon, Zoran Popovi, David Baker, and Foldit Players. Algorithm discovery by protein folding game players. Proceedings of the National Academy of Sciences, 108(47):1894918953, November 2011.

[31] Jack B. Maguire, Hugh K. Haddox, Devin Strickland, et al. Perturbing the energy land-scape for improved packing during computational protein design. Proteins: Structure, Function, and Bioinformatics, 89(4):436449, December 2020.

